# Reference ranges for cardiovascular function during exercise: effects of aging and gender on performance

**DOI:** 10.1101/2023.08.23.23294458

**Authors:** Ronny Schweitzer, Antonio de Marvao, Mit Shah, Paolo Inglese, Peter Kellman, Alaine Berry, Ben Statton, Declan P O’Regan

## Abstract

**Purpose:** Real-time (RT) exercise cardiac magnetic resonance imaging (exCMR) provides a highly reproducible and accurate assessment of cardiac volumes during maximal exercise. It has advantages over alternative approaches due to its high spatial resolution and use of physiological stress. Here we define the healthy response to exercise in adults and the effects of age and gender on performance.

**Materials and Methods:** Between 2018 and 2021, we conducted CMR evaluation on 169 healthy adults who had no known cardiovascular disease, did not harbour genetic variants associated with cardiomyopathy, and who completed an exCMR protocol using a pedal ergometer. Participants were imaged at rest and after exercise with left ventricular parameters measured using commercial software by two readers. Eight participants were excluded from the final analysis due to poor image quality and/or technical issues. Prediction intervals were calculated for each parameter.

**Results:** Exercise caused an increase in heart rate (64±9 bpm vs 133±19 bpm, *P* < 0.001), left ventricular end-diastolic volume (140±32 ml vs 148±35 ml, *P* < 0.001), stroke volume (82±18 ml vs 102±25 ml, *P* < 0.001), ejection fraction (59±6% vs 69±7%, *P* < 0.001), and cardiac output (5.2±1.1 l/min vs 13.5±3.9 l/min, *P* < 0.001), with a decrease in left ventricular end-systolic volume (58±18 ml vs 46±15 ml, *P* < 0.001). There was an effect of gender and age on response to exercise across most parameters. Measurements showed good to excellent intra- and inter-observer agreement.

**Conclusion:** In healthy adults, an increase in cardiac output after exercise is driven by a rise in heart rate with both increased ventricular filling and emptying. We establish normal ranges for exercise response, stratified by age and gender, as a reference for the use of exCMR in clinical practice.

## Introduction

Exercise cardiac magnetic resonance imaging (exCMR) is a non-invasive imaging modality that offers high image quality during physiological stress. It can be undertaken as part of a comprehensive CMR study of structure, function and tissue characterisation to assess physiological reserve during maximal exercise. This is typically performed using real-time cine imaging, as well as other CMR sequences to assess myocardial strain, aortic flow or tissue metabolism. The technique has emerging applications across several conditions including the stratification of patients with pulmonary hypertension, diagnosing athlete’s heart, evaluating exercise tolerance in congenital heart disease and in the detection of ischemia.^1^ The use of exCMR offers significant advantages over exercise echocardiography in its reliable assessment of systolic and diastolic function, which is less user-dependent.^2^ There is also no requirement for breath-holding, which may itself affect cardiac filling pressures.^3^ Physical exercise has a higher diagnostic sensitivity and fewer adverse events compared to pharmacological stressors and is recommended for stress imaging whenever feasible.^4^

Knowledge of the range of normal function in exercise is needed for clinical interpretation,^5^ but to date there are no reported reference ranges for exCMR. With widening use of exCMR in both clinical and research domains, there is a need to develop representative values in healthy individuals. In this study, we report findings from exCMR in a cohort of healthy adults, with no history of cardiovascular disease or major comorbidities. Participants were also screened for rare pathogenic or likely pathogenic genetic variants associated with cardiomyopathy to ensure there were no sub-clinical cases of inherited cardiovascular disease. Imaging was performed using contemporary protocols, exercise regimes and analysis methodology to develop age and gender stratified normal ranges to aid in the interpretation of exCMR.

## Materials and Methods

### Study Design and Population

This was a prospective, observational, single centre study performed at Imperial College London. The study was approved by the National Research Ethics Service (17/LO/0034) and all participants provided written informed consent. The overall study aimed to evaluate the role of genetic variation on cardiovascular function in a clinically healthy cohort using CMR imaging. The data presented are for a sub-study that used exCMR to assess functional reserve.

Enrollment was between January 2018 and April 2021. Participants were eligible for inclusion if they were over the age of 18 years, had no history of cardiovascular disease, and were genotype-negative for inherited cardiac conditions during genetic screening. Exclusion criteria included known or suspected pregnancy, physical limitations that would preclude participants from exercising, and any contraindications to MRI. All participants completed a health questionnaire and underwent assessment of height and weight, blood pressure, and a resting 12-lead electrocardiogram (ECG).

### Genetic Sequencing

Genetic sequencing of 169 genes, associated with inherited cardiac conditions, was conducted using the Illumina MiSeq, Illumina NextSeq, or Life Technologies SOLiD 5500xl platforms, after target enrichment using in-solution hybridisation (Illumina Nextera or Agilent SureSelect), as previously described.^6–8^ Variants were annotated and filtered using a validated bioinformatics pipeline.^9^ Rare variants were defined by a minor allele frequency of <10^-4^ from population data in gnomAD applying stringent filtering. Individuals harbouring variants that would be called pathogenic or likely pathogenic if identified in a patient with an inherited cardiac condition were considered genotype-positive.

### Image Acquisition

Images were acquired using a 1.5T MAGNETOM Aera (Siemens Healthineers, Erlangen, Germany) using a 60 channel cardiac coil. Participants underwent a complete CMR protocol including structural and functional imaging, phase-contrast flow assessment and late-gadolinium enhancement (Figure 1) following published guidelines and standards.^10^ Following this, short axis cines covering the entire left ventricle were acquired at rest and exercise using a real time (RT) balanced steady-state free precession (bSSFP) sequence. Thirteen RT short-axis slices were positioned to give complete coverage from base to apex allowing for potential patient movement during exercise. Each slice was triggered on the ECG R-wave and then acquired free-running for 3 seconds to acquire 90 frames. A single cardiac cycle of 30 frames at each slice location was reconstructed retrospectively using interpolation. Typical acquisition parameters were: TR = 2.2 ms, TE = 0.97 ms, slice thickness = 8 mm, slice gap = 2 mm, matrix size = 128 x 96, field of view = 360 x 270 mm. GeneRalized Autocalibrating Partial Parallel Acquisition (GRAPPA) acceleration (factor = 4) was used achieving a temporal resolution of 31 ms.

**Figure 1.**
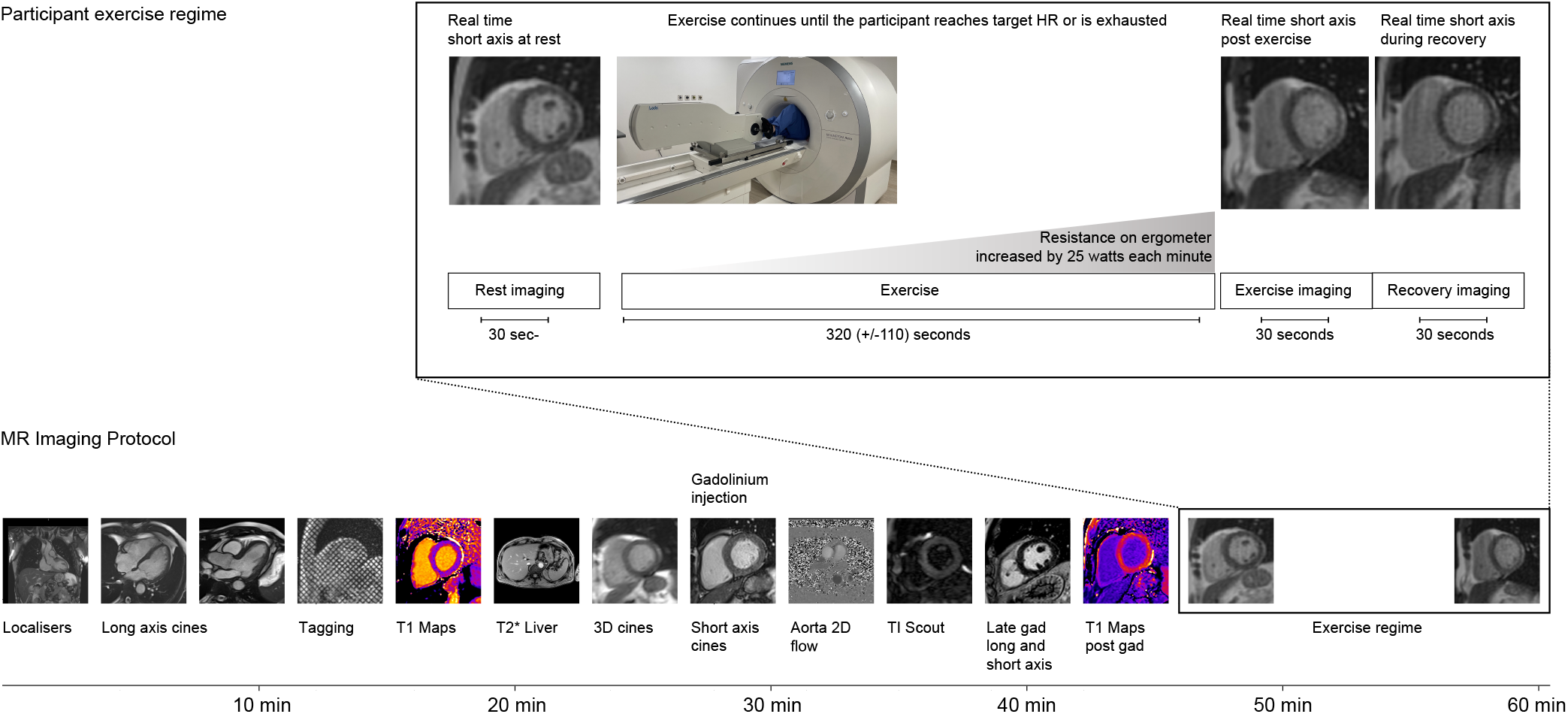
Imaging protocol. Exercise regime and imaging protocol used in the study.

### Exercise Protocol

Exercise was performed using a MR-conditional variable resistance supine ergometer (Lode BV, Netherlands; Figure 2) attached to the scanner table. Participants were asked to grip the handles of the ergometer and coached to keep their torso as stationary as possible during exercise to minimize bulk movement. The target heart rate (HR) for each participant was calculated as 85% of the maximal HR using Fox’s formula (220 - age in years).^11^

**Figure 2.**
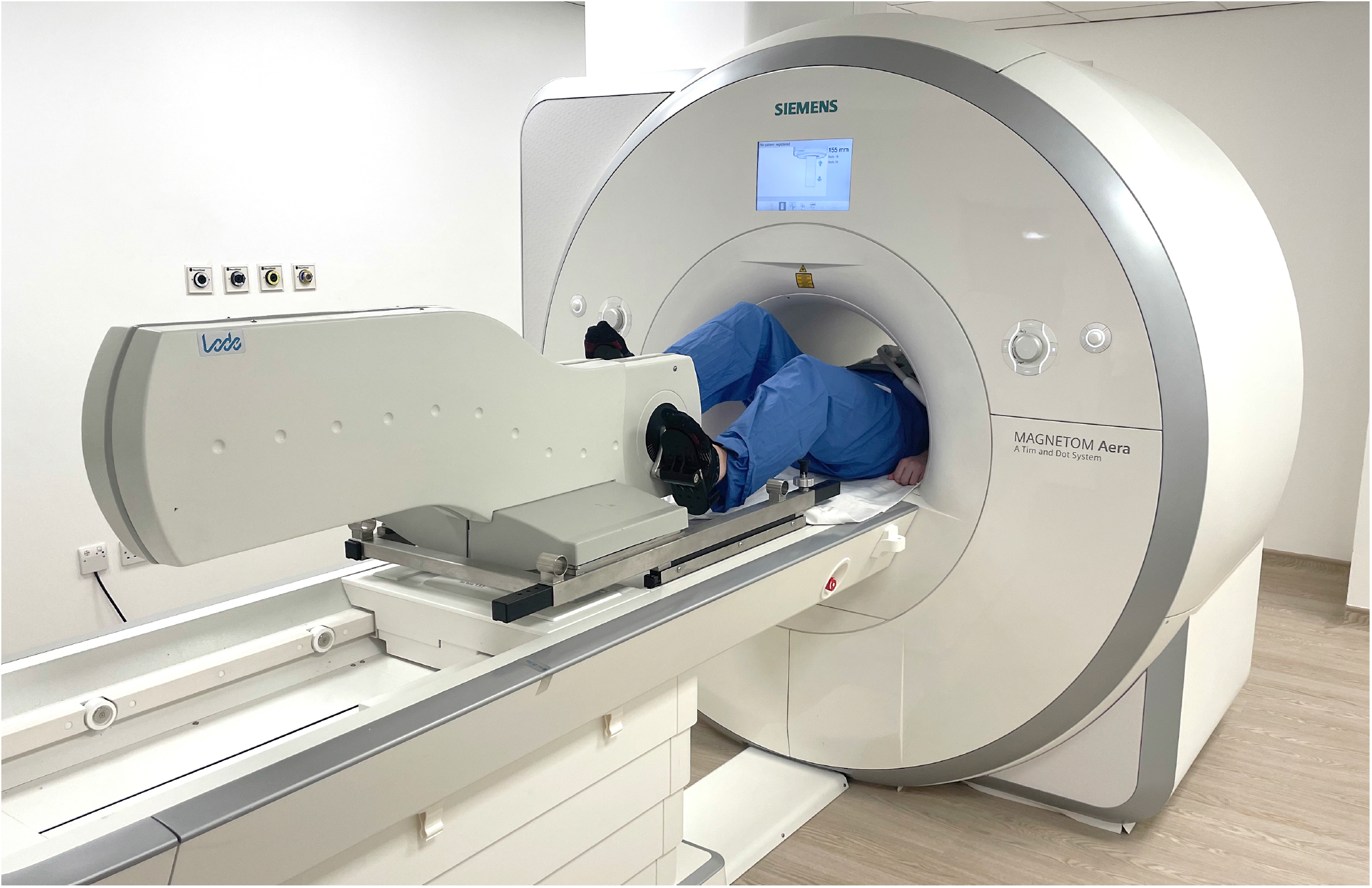
Pedal ergometer. Positioning of the bed-mounted MR-conditional pedal ergometer allowing participants to exercise while at isocentre (hand holds not shown). An electro-magnetic braking system allows a maximum peak load of up to 300W. An external control unit displays multiple ergometry parameters in real time.

Following resting RT imaging, the participants were instructed to start exercising while maintaining a cadence of 70 - 80 rpm, with an initial minimal workload for 1 minute. While the participants continued to exercise in the bore of the MR scanner, the workload was increased by 25 W every minute until the target HR was reached or leg exhaustion occurred. The free-breathing RT imaging began immediately following the cessation of exercise in order to minimise movement artefacts and ECG mis-triggering.

### Image Analysis

In this study only breath-hold cine images and free-breathing RT images were analysed. All images were analysed using cvi42 (Circle Cardiovascular Imaging Inc., Calgary, Canada, version 5.16). End-systolic and end-diastolic frames were manually selected. Subsequently, left ventricular endo- and epicardial borders were traced using automated deep learning segmentation^12^ and were then visually inspected and corrected if necessary (Figure 3). Papillary muscles were included in the left ventricular blood pool. Absolute values were indexed to body surface area (BSA) using the DuBois method. Cardiac output and cardiac index were calculated using the maximal heart rate achieved.

**Figure 3.**
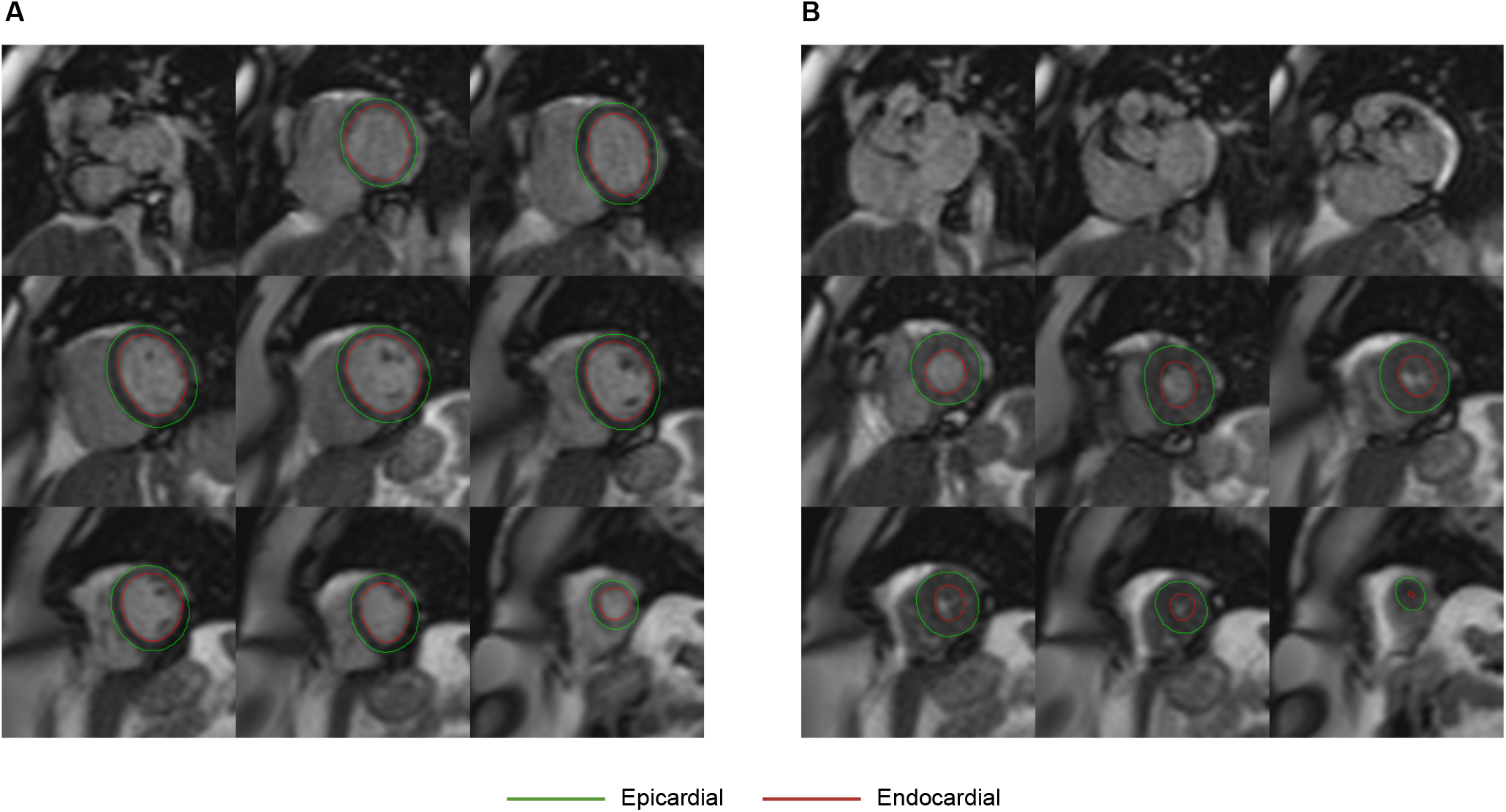
Real-time cine imaging. Example of exercise real-time short axis cine images from one participant at a) end diastole and b) end systole. Epicardial contours are shown in green and the endocardial contours in red.

### Intra- and Inter-observer Reliability

Inter- and intra-observer variability were evaluated to assess reliability of the CMR measurements. Fifty participants were randomly selected and re-analyzed after an interval of 3 months by two operators blinded to each other’s analysis.

### Statistical Analysis

Statistical analyses were performed using R (version 4.2.1, R Core Team, Vienna, Austria) and Python (version 3.8.5, Python Software Foundation, Delaware, United States). Variables were expressed as percentages, if categorical, and as mean ± standard deviation (SD), if continuous. Bland-Altman plots were used to quantify the agreement between RT and breath-hold cines at rest. Intra-class correlation coefficient (ICC) was used to assess reliability and categorized as poor (<0.5), moderate (0.5-0.75), good (0.75-0.9), and excellent (>0.9).^13^ Reference ranges were defined as the 95% prediction interval of the Student’s t-distribution. Ranges were stratified by gender and age groups (20-39, 40-59, 60-79 years). Comparisons were adjusted for age unless stratified by age group. Paired t-tests were used to compare rest and exercise, unpaired t-tests to compare genders, and one-way analysis of variance (ANOVA) to compare age groups. Post hoc testing used Tukey’s honest significant difference test (THSD). The t-tests were two-sided and the ANOVA one-sided. There was no adjustment for multiple comparisons. A *P* value ≤ 0.05 was considered significant.

## Results

A total of 177 healthy, genotype-negative volunteers were enrolled, of whom 161 were included in the sub-study (Figure 4). The mean age was 49±14 years (range 22-77) with 70% of participants being Caucasian and 53% women. Baseline characteristics are shown in Table 1. The RT images were typically acquired over 27 seconds to achieve coverage of the heart from base to apex during which time there was a mean difference of 12 beats per minute from peak heart rate.

**Table 1.** Participant characteristics. Demographic and physiological parameters for the study participants. Heart rate shown at rest and peak exercise. BSA, body surface area; BMI, body mass index; bpm, beats per minute; HR, heart rate.

|  | 20 - 39 years<br>n = 50 | 40 - 59 years<br>n = 67 | 60 - 79 years<br>n = 44 |
| --- | --- | --- | --- |
| Age (years) | 32 ± 4 | 51 ± 5 | 67 ± 4 |
| Men (%) | 52.0 | 47.8 | 40.9 |
| Caucasian (%) | 56.0 | 72.7 | 82.9 |
| Weight (kg) | 75 ± 14 | 78 ± 17 | 72 ± 16 |
| Height (cm) | 173 ± 9 | 171 ± 10 | 168 ± 10 |
| BSA (m<sup>2</sup>) | 1.89 ± 0.21 | 1.9 ± 0.23 | 1.82 ± 0.23 |
| BMI (kg/m<sup>2</sup>) | 24.98 ± 3.88 | 26.78 ± 5.11 | 25.28 ± 4.67 |
| Systolic Blood Pressure (mmHg) | 122 ± 13 | 128 ± 14 | 136 ± 20 |
| Diastolic Blood Pressure (mmHg) | 75 ± 8 | 81 ± 8 | 80 ± 10 |
| HR at rest (beats per minute) | 64 ± 7 | 65 ± 10 | 63 ± 7 |
| HR during exercise (beats per minute) | 145 ± 17 | 130 ± 16 | 122 ± 15 |
| Goal HR reached? (%) | 38.0 | 31.3 | 47.7 |
| Wattage Max (W) | 141 ± 34 | 117 ± 38 | 101 ± 36 |

**Figure 4.**
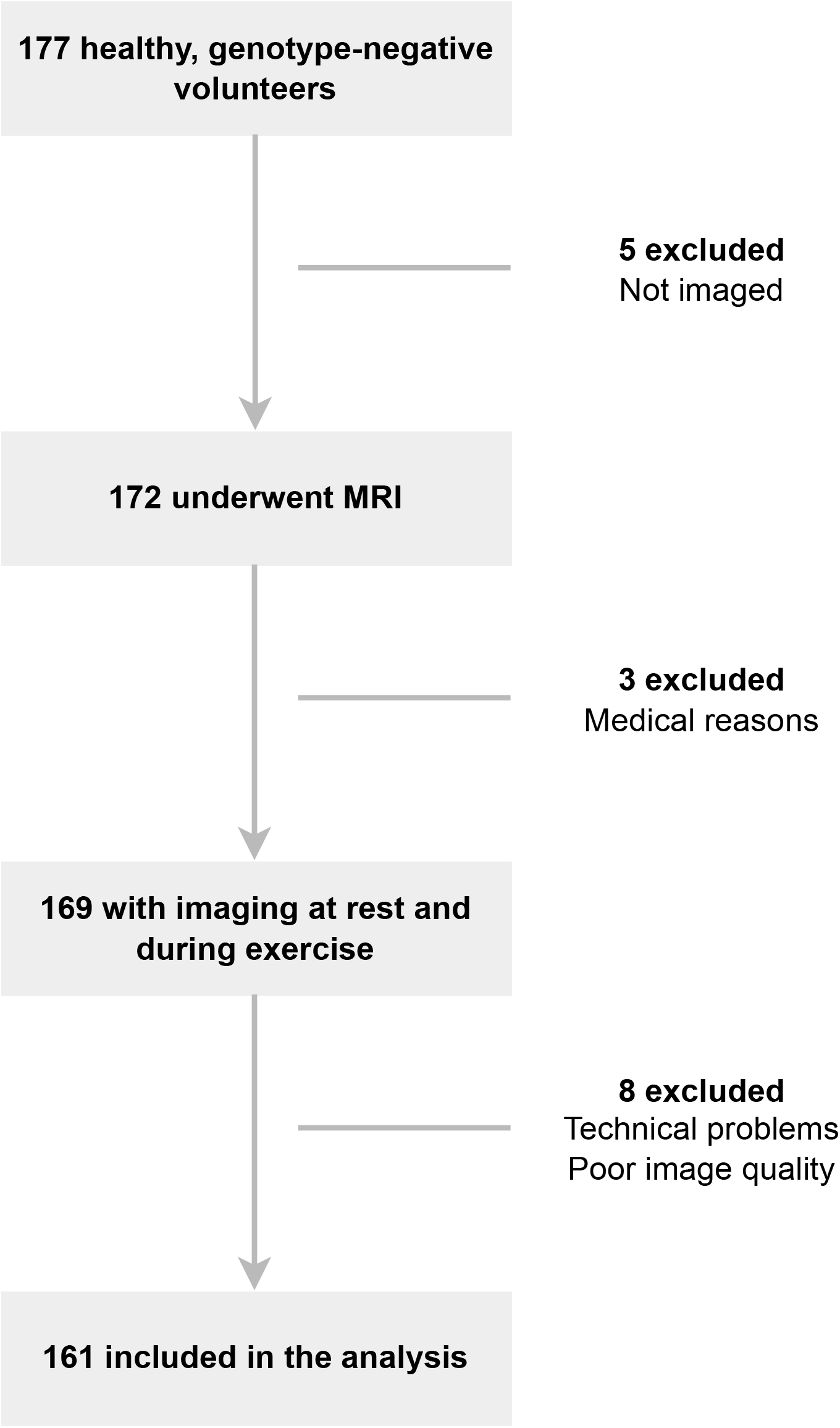
Study flowchart. MRI, magnetic resonance imaging.

### Left Ventricular Reference Values

Reference ranges for absolute and relative change in left ventricular parameters during exercise are stratified by gender and age (men: Table 2, women: Table 3). The “lower” value represents the lower end of the 95% prediction interval, the “mean” value represents the average change, and the “upper” value represents the upper end of the 95% prediction interval.

**Table 2.** Absolute and relative reference ranges on exercise by age group in men. EDV, end-diastolic volume; ESV, end-systolic volume; SV, stroke volume; EF, ejection fraction; CO, cardiac output; CI, cardiac index; BSA, body surface area.

|  | 20 - 39 years |  |  | 40 - 59 years |  |  | 60 - 79 years |  |  |
| --- | --- | --- | --- | --- | --- | --- | --- | --- | --- |
|  | lower | mean | upper | lower | mean | upper | lower | mean | upper |
| EDV - ml (% change) | -10 (-5%) | 0 (0%) | 9 (6%) | 11 (6%) | 18 (11%) | 24 (16%) | 0 (0%) | 11 (7%) | 21 (14%) |
| EDV/BSA - ml/m<sup>2</sup> (% change) | -5 (-5%) | 0 (0%) | 5 (6%) | 5 (6%) | 9 (11%) | 12 (16%) | 0 (0%) | 5 (7%) | 10 (14%) |
| ESV - ml (% change) | -26 (-34%) | -21 (-27%) | -15 (-19%) | -20 (-27%) | -15 (-20%) | -11 (-14%) | -18 (-28%) | -11 (-15%) | -3 (-2%) |
| ESV/BSA - ml/m<sup>2</sup> (% change) | -13 (-34%) | -10 (-27%) | -8 (-19%) | -9 (-27%) | -7 (-20%) | -5 (-14%) | -9 (-28%) | -5 (-15%) | -2 (-2%) |
| SV - ml (% change) | 11 (13%) | 20 (23%) | 29 (34%) | 26 (30%) | 33 (39%) | 40 (47%) | 14 (15%) | 22 (24%) | 29 (33%) |
| SV/BSA - ml/m<sup>2</sup> (% change) | 6 (13%) | 10 (23%) | 15 (34%) | 13 (30%) | 16 (39%) | 19 (47%) | 7 (15%) | 10 (24%) | 14 (33%) |
| EF - % (% change) | 9 (16%) | 12 (22%) | 15 (28%) | 11 (19%) | 13 (24%) | 15 (29%) | 6 (9%) | 9 (16%) | 13 (22%) |
| CO - l/min (% change) | 9.4 (166%) | 10.9 (203%) | 12.4 (240%) | 8.6 (153%) | 10.0 (185%) | 11.5 (217%) | 7.5 (129%) | 8.5 (150%) | 9.5 (170%) |
| CI - l/min/m<sup>2</sup> (% change) | 4.7 (166%) | 5.5 (203%) | 6.2 (240%) | 4.1 (153%) | 4.8 (185%) | 5.5 (217%) | 3.6 (129%) | 4.2 (150%) | 4.8 (170%) |

**Table 3.** Absolute and relative reference ranges on exercise by age group in women. EDV, end-diastolic volume; ESV, end-systolic volume; SV, stroke volume; EF, ejection fraction; CO, cardiac output; CI, cardiac index; BSA, body surface area.

|  | 20 - 39 years |  |  | 40 - 59 years |  |  | 60 - 79 years |  |  |
| --- | --- | --- | --- | --- | --- | --- | --- | --- | --- |
|  | lower | mean | upper | lower | mean | upper | lower | mean | upper |
| EDV - ml (% change) | -1 (0%) | 4 (3%) | 11 (8%) | -1 (0%) | 4 (5%) | 11 (10%) | 2 (3%) | 9 (10%) | 16 (18%) |
| EDV/BSA - ml/m<sup>2</sup> (% change) | 0 (0%) | 2 (3%) | 6 (8%) | 0 (0%) | 2 (5%) | 6 (10%) | 1 (3%) | 5 (10%) | 9 (18%) |
| ESV - ml (% change) | -16 (-31%) | -12 (-21%) | -8 (-12%) | -10 (-23%) | -7 (-16%) | -4 (-9%) | -7 (-15%) | -4 (-7%) | 0 (0%) |
| ESV/BSA - ml/m<sup>2</sup> (% change) | -9 (-31%) | -7 (-21%) | -4 (-12%) | -5 (-23%) | -4 (-16%) | -2 (-9%) | -4 (-15%) | -2 (-7%) | 0 (0%) |
| SV - ml (% change) | 10 (14%) | 17 (21%) | 24 (29%) | 5 (11%) | 12 (20%) | 19 (30%) | 6 (12%) | 13 (24%) | 21 (36%) |
| SV/BSA - ml/m<sup>2</sup> (% change) | 6 (14%) | 9 (21%) | 13 (29%) | 3 (11%) | 7 (20%) | 10 (30%) | 3 (12%) | 8 (24%) | 12 (36%) |
| EF - % (% change) | 7 (12%) | 10 (17%) | 13 (22%) | 5 (9%) | 8 (13%) | 10 (18%) | 2 (5%) | 6 (11%) | 9 (16%) |
| CO - l/min (% change) | 7.0 (136%) | 8.3 (161%) | 9.7 (185%) | 5.9 (125%) | 6.6 (144%) | 7.4 (162%) | 4.9 (110%) | 5.7 (133%) | 6.5 (155%) |
| CI - l/min/m<sup>2</sup> (% change) | 4.0 (136%) | 4.7 (161%) | 5.4 (185%) | 3.3 (125%) | 3.7 (144%) | 4.2 (162%) | 2.9 (110%) | 3.4 (133%) | 3.8 (155%) |

### Left Ventricular Response to Exercise

For the cohort as a whole, exercise caused an increase in HR (64±9 bpm vs 133±19 bpm, *P* < 0.001), stroke volume (SV) (82±18 ml vs 102±25 ml, *P* < 0.001), ejection fraction (EF) (59±6% vs 69±7%, *P* < 0.001), and cardiac output (CO) (5.2±1.1 l/min vs 13.5±3.9 l/min, *P* < 0.001). The increase in EF and SV was predominantly mediated by a reduction in end-systolic volume (ESV) (58±18 ml vs 46±15 ml, *P* < 0.001), with a smaller increase in end-diastolic volume (EDV) (140±32 ml vs 148±35 ml, *P* < 0.001).

### Gender Differences

Comparisons between left ventricular parameters at rest and exercise are shown in Table 4 and Table S1. Stratified plots of participant-level response to exercise are shown in Figure 5, Figure 6, and Figure S1. In summary, men had higher resting indexed EDV (79±12 ml/m^2^ vs 70±13 ml/m^2^, *P* < 0.001) and ESV (34±7 ml/m^2^ vs 27±7 ml/m^2^, *P* < 0.001) than women, and although HR increase was similar (72±22 bpm vs 65±18 bpm, *P* = 0.105), men achieved a higher indexed SV (57±11 ml/m^2^ vs 51±9 ml/m^2^, *P* < 0.001) through greater left ventricular emptying (change in indexed ESV: -8±7 ml/m^2^ vs -4±5 ml/m^2^, *P* = 0.002).

**Table 4.** Comparison of rest vs exercise in both genders. Values adjusted for age. EDV, end-diastolic volume; ESV, end-systolic volume; SV, stroke volume; EF, ejection fraction; CO, cardiac output; CI, cardiac index; BSA, body surface area; bpm, beats per minute.

|  | Men |  |  | Women |  |  |
| --- | --- | --- | --- | --- | --- | --- |
|  | Rest<br>n = 76 | Exercise<br>n = 76 | P value | Rest<br>n = 85 | Exercise<br>n = 85 | P value |
| HR (bpm) | 64 ± 11 | 135 ± 21 | <0.001 | 65 ± 7 | 130 ± 16 | <0.001 |
| EDV (ml) | 160 ± 27 | 170 ± 32 | <0.001 | 122 ± 25 | 129 ± 25 | 0.024 |
| EDV/BSA (ml/m<sup>2</sup>) | 79 ± 12 | 83 ± 14 | <0.001 | 70 ± 13 | 73 ± 12 | 0.015 |
| ESV (ml) | 70 ± 16 | 54 ± 15 | <0.001 | 47 ± 12 | 39 ± 12 | <0.001 |
| ESV/BSA (ml/m<sup>2</sup>) | 34 ± 7 | 26 ± 7 | <0.001 | 27 ± 7 | 22 ± 7 | <0.001 |
| SV (ml) | 90 ± 16 | 116 ± 24 | <0.001 | 75 ± 16 | 89 ± 19 | <0.001 |
| SV/BSA (ml/m<sup>2</sup>) | 44 ± 8 | 57 ± 11 | <0.001 | 43 ± 9 | 51 ± 9 | <0.001 |
| EF (%) | 56 ± 6 | 68 ± 7 | <0.001 | 61 ± 6 | 70 ± 7 | <0.001 |
| CO (l/min) | 5.7 ± 1.1 | 15.6 ± 3.7 | <0.001 | 4.8 ± 1.1 | 11.7 ± 3.0 | <0.001 |
| CI (l/min/m<sup>2</sup>) | 2.8 ± 0.5 | 7.7 ± 1.7 | <0.001 | 2.8 ± 0.5 | 6.6 ± 1.6 | <0.001 |

**Figure 5.**
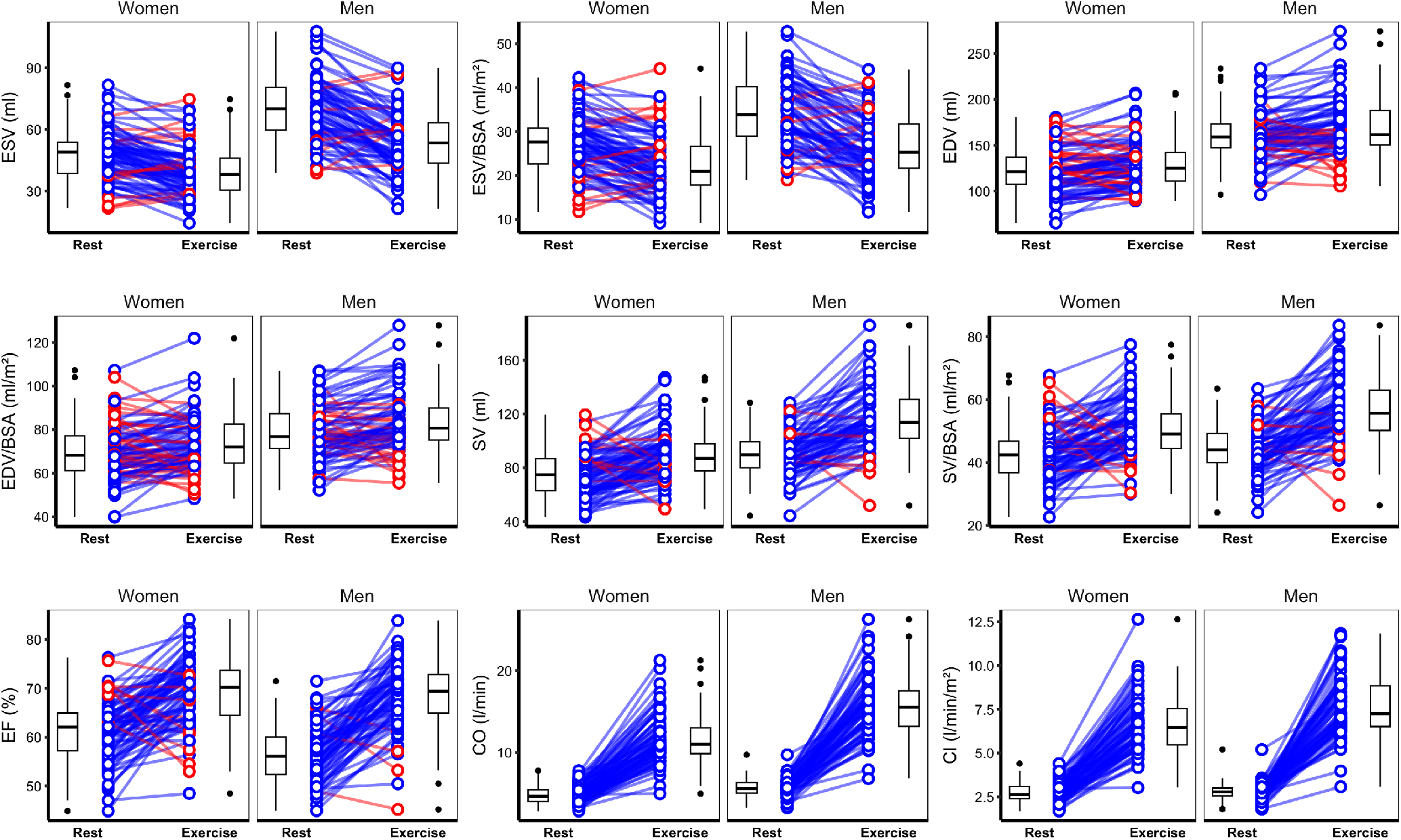
Left ventricular parameters at rest and exercise stratified by gender. Lines join corresponding values. Boxes show median and inter-quartile range (IQR), and whiskers 1.5*IQR. Individual direction of response shown in blue (same) or red (different) with respect to average change. EDV, end-diastolic volume; ESV, end-systolic volume; SV, stroke volume; EF, ejection fraction; CO, cardiac output; CI, cardiac index; BSA, body surface area. n = 161.

**Figure 6.**
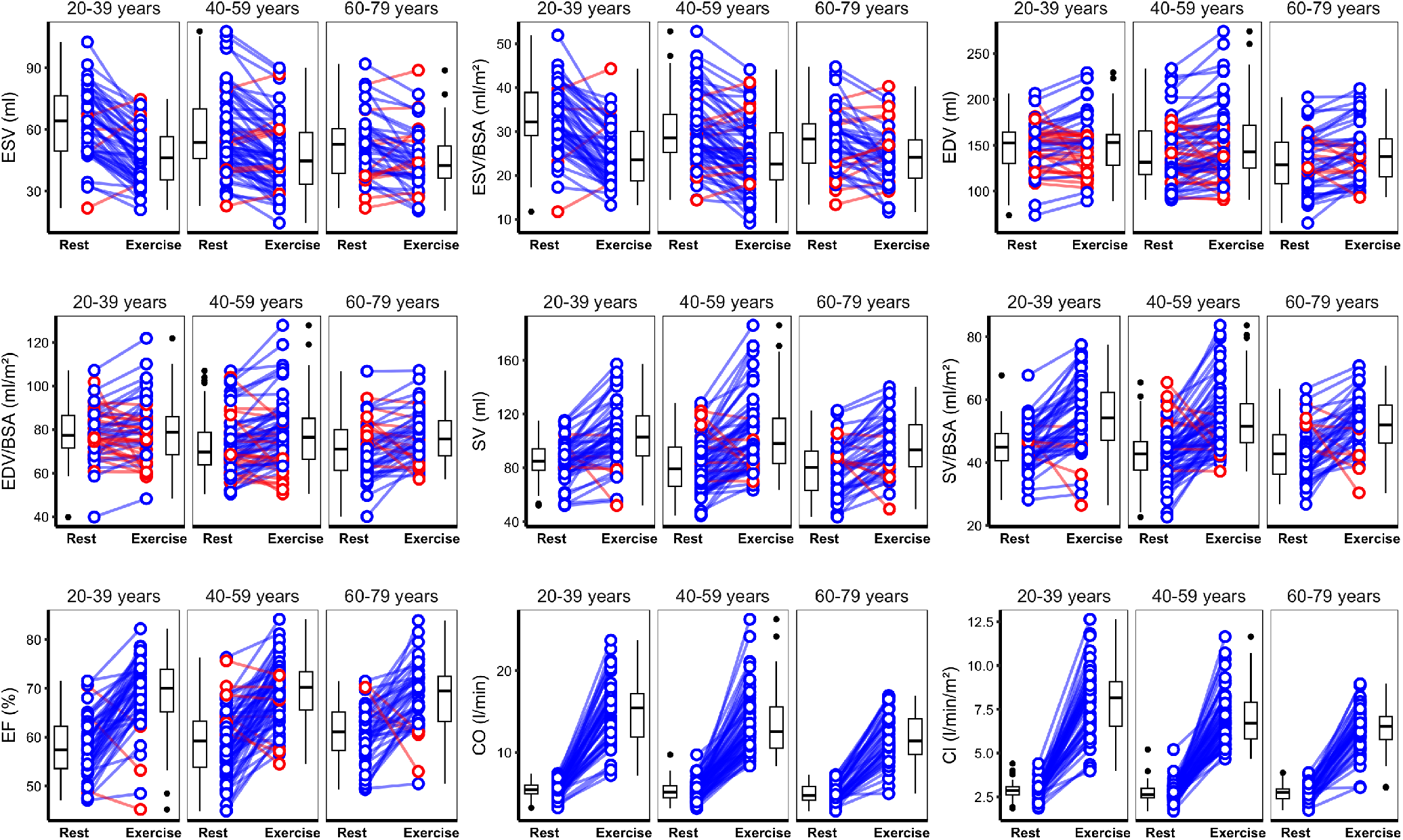
Left ventricular parameters at rest and exercise stratified by age group. Lines join corresponding values. Boxes show median and inter-quartile range (IQR), and whiskers 1.5*IQR. Individual direction of response shown in blue (same) or red (different) with respect to average change. EDV, end-diastolic volume; ESV, end-systolic volume; SV, stroke volume; EF, ejection fraction; CO, cardiac output; CI, cardiac index; BSA, body surface area. n = 161.

### Age Group Differences

Comparisons between left ventricular parameters at rest and exercise stratified by gender and age group are shown in Tables S3 to S6. In summary, HR response to exercise declined with age in both genders (men and women: *P* < 0.01). Older adults showed less ventricular emptying (change in indexed ESV men: *P* = 0.042; women: *P* = 0.006) but men achieved an increase in indexed EDV (men: *P* = 0.01; women: *P* = 0.454), maintaining a similar exercise EF (men and women: *P* > 0.05) with a tendency to an increase in CI across age groups (men: *P* = 0.059; women: *P* = <0.001).

### Data Reliability

Bland-Altman plots (Figure S2) comparing resting RT imaging with breath-hold cines showed a good level of agreement and no measurement bias. Reliability testing demonstrated good to excellent agreement for all measured variables (Table S7).

## Discussion

This study reports the effects of exercise in healthy volunteers assessed with exCMR and provides corresponding reference ranges stratified by age and gender. Defining normal ranges for left ventricular parameters will enable exCMR findings to be placed in the context of healthy variation and will facilitate non-pharmacological, quantitative assessment of functional reserve. The availability of this normative data will assist with the clinical interpretation of exCMR as it becomes more widely used in both research settings and clinical practice.

CMR is considered the gold standard for assessing resting cardiac function and a single exam provides quantitative information about cardiac structure, tissue characteristics and blood flow.^14^ CMR has multiple Class I and Class 2a recommendations for pharmacological functional testing,^15^ and more recently exCMR has emerged as a non-pharmacological assessment of physiological reserve that is safe, accurate, and cost effective.^1,16^ The EXACT trial showed the potential of using treadmill exCMR in patients at risk for coronary artery disease, demonstrating a strong agreement with coronary angiography and outperforming SPECT.^17^ Additionally, the EMPIRE study showed excellent agreement between supine exCMR and invasive coronary fractional flow reserve.^18^ The use of exCMR also provides a non-invasive approach to identify heart failure with preserved ejection fraction in cases where both clinical and echocardiographic findings are inconclusive.^19^ Further applications of exCMR include differentiating between athlete’s heart and dilated cardiomyopathy,^20,21^ as well as stratification of patients with pulmonary hypertension.^22^

The widening availability and indications for exCMR highlight the need for published normal ranges which have not previously been reported for this technique.^5^ There are variations in practice regarding exercise regimens, imaging protocols and post-processing as this is still an emerging technique. For the study population, we recruited healthy volunteers in the UK that had been pre-screened for cardiovascular disease, including incidental cardiomyopathy-associated rare variants. The population had an age range from 21 to 77 and had equal numbers of men and women, but was 70% Caucasian. Analyses were stratified by age and gender. We did not have a sufficient sample size to stratify by ancestry. The ranges therefore represent a community-based urban reference population free of known disease. The exercise regimen is based on reaching an age predicted maximal HR to individualise exercise intensity,^23^ although some studies have used upright cardiopulmonary exercise testing (CPET) to adjust target power output,^22,24^ or used a single target HR.^2^ Our protocol has the practical advantage of being personalised while not requiring pre-test CPET. There are also a variety of exercise equipment and protocols available including treadmill exercise outside the magnet. While MR conditional cycle ergometers are expensive and currently less widely available, they offer a means of accurately controlling exercise intensity while inside the bore of the magnet with immediate imaging reducing potential sources of variability. Of those that underwent exCMR only 5% were excluded from subsequent analysis. Imaging protocols for exCMR favour RT imaging as the patient is free-breathing after exercise. While accelerated RT sequences continue to be developed to improve temporal resolution and mitigate motion artefact,^25^ we used a product sequence that is widely available, shows strong agreement with gated cine sequences and achieves good reproducibility. To facilitate analysis of end-diastolic and end-systolic frames on RT imaging, we used a reconstruction that produces a consistent number of frames, but this can also be readily achieved without interpolation. For post-processing we used widely available commercial software for automated assessment initially followed by manual refinement. The overall protocol is broadly generalisable and reflects current practice.

A consistent response to exercise has been shown in meta-analyses of exCMR across different field strengths, exercise protocols, and imaging techniques.^26^ Our cohort demonstrated a robust response to supine exercise with almost a 250% increase in cardiac output achieving at least the maximal response to similar protocols.^20^ Overall, EDV showed only a modest and variable increase on exercise, while there was a significant rise in HR and decrease in ESV. Older adults, with a reduced HR response and less systolic reserve, relied on greater ventricular filling to maintain exercise CO. Higher HR in younger adults during exercise, with decreased filling time, could account for the limited increase in EDV during physical activity.^27^ In the oldest age groups, a decline in diastolic relaxation could also contribute to a blunted rise in CO.^28^ Men had higher indexed resting and exercise volumes than women and a greater absolute and relative response to exercise.^29^ The rise in HR was similar between genders, and a higher exercise SV in men was achieved through greater left ventricular emptying. We also demonstrated that it was feasible to incorporate exCMR into a full clinical protocol, including tissue characterisation and late enhancement, that could be completed within 60 minutes with a low technical failure rate. Although confined to healthy volunteers, this shows how exCMR could be incorporated into routine CMR practice.

### Limitations

Our cohort mainly comprised Caucasians, which could limit the generalizability of our reference ranges to other ancestries. Baseline fitness and lifestyle can also significantly affect cardiac function even in non-athletes.^30^ The study was conducted at a single center and we did not assess the variability due to other scanners or protocols. We used widely available sequences and post-processing, but novel sequences to increase temporal and spatial resolution could potentially achieve better reliability and functional characterisation.^31^ We did not assess test-retest performance due to the effects of fatigue.

### Conclusions

This study establishes the normal response to exercise assessed by exCMR and provides left ventricular adult reference ranges in a community-based cohort. Quantitative evaluation of exCMR using these normative data will enable better differentiation between health and disease in clinical practice.

## Data Availability

The code used to pre-process and analyse the data, as well as aggregated exCMR parameters, is available at https://github.com/ImperialCollegeLondon/exCMR (doi: 10.5281/zenodo.10301641).

## Availability of data and materials

Data generated or analyzed during the study are available from the corresponding author by request. The code used to pre-process and analyse the data, as well as aggregated exCMR parameters, is available at https://github.com/ImperialCollegeLondon/exCMR (doi: 10.5281/zenodo.10301641).

### Funding

The study was supported by the Medical Research Council (MC_UP_1605/13); the British Heart Foundation (RG/19/6/34387, RE/18/4/34215); and the National Institute for Health Research (NIHR) Imperial College Biomedical Research Centre. For the purpose of open access, the authors have applied a Creative Commons Attribution (CC BY) licence to any author accepted manuscript version arising.

## Acknowledgements

We thank our research radiographer Marina Quinlan, and our senior clinical research nurse Maria Brezitski-Abramova (Imperial College London) for their contribution to this work.

## Supplementary Figures

**Figure S1.**
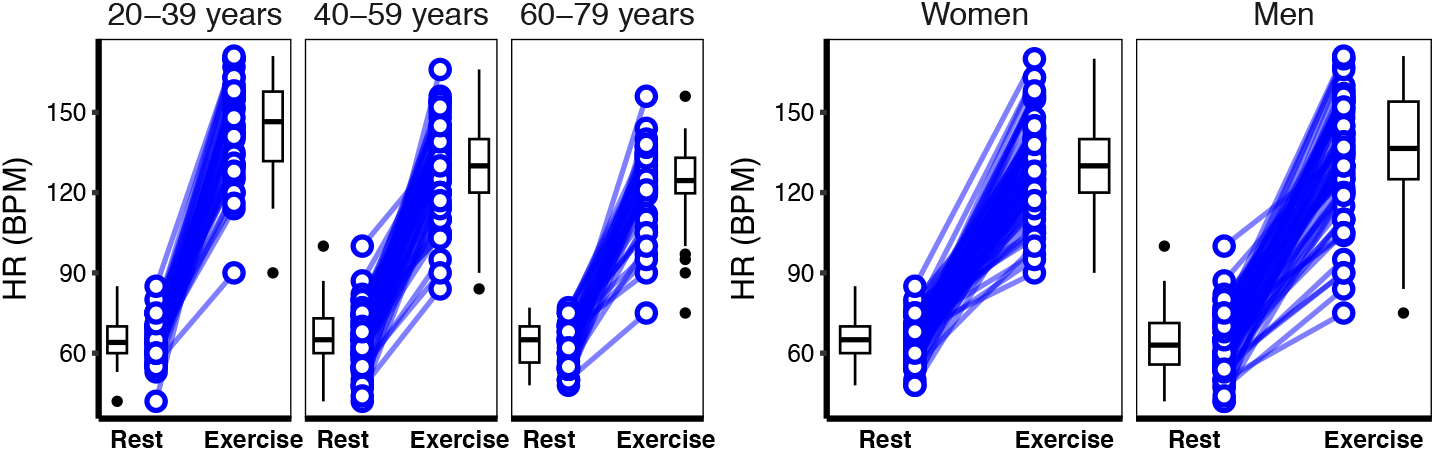
Heart rate at rest and exercise stratified by age and gender. Boxes show median and inter-quartile range (IQR), and whiskers 1.5*IQR. HR, heart rate; BPM, beats per minute. n = 161.

**Figure S2.**
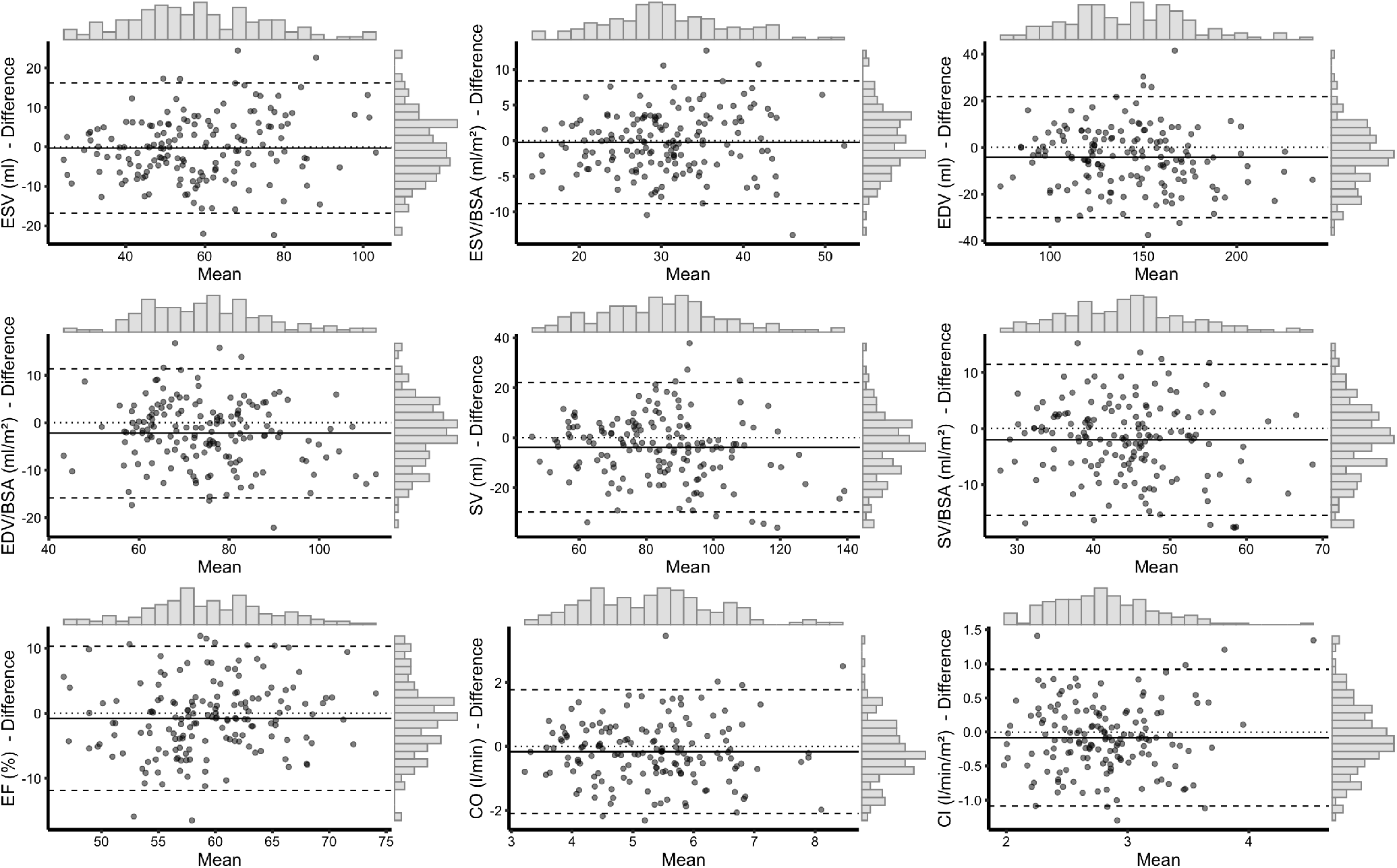
Agreement between methods to assess resting function. Bland-Altman plots with marginal histograms comparing left ventricular parameters derived from retrospectively gated cine images to real time cine images at rest. RT, real time; EDV, end-diastolic volume; ESV, end-systolic volume; SV, stroke volume; EF, ejection fraction; CO, cardiac output; CI cardiac index; BSA, body surface area. n = 161.

## Supplementary Tables

**Table S1.**
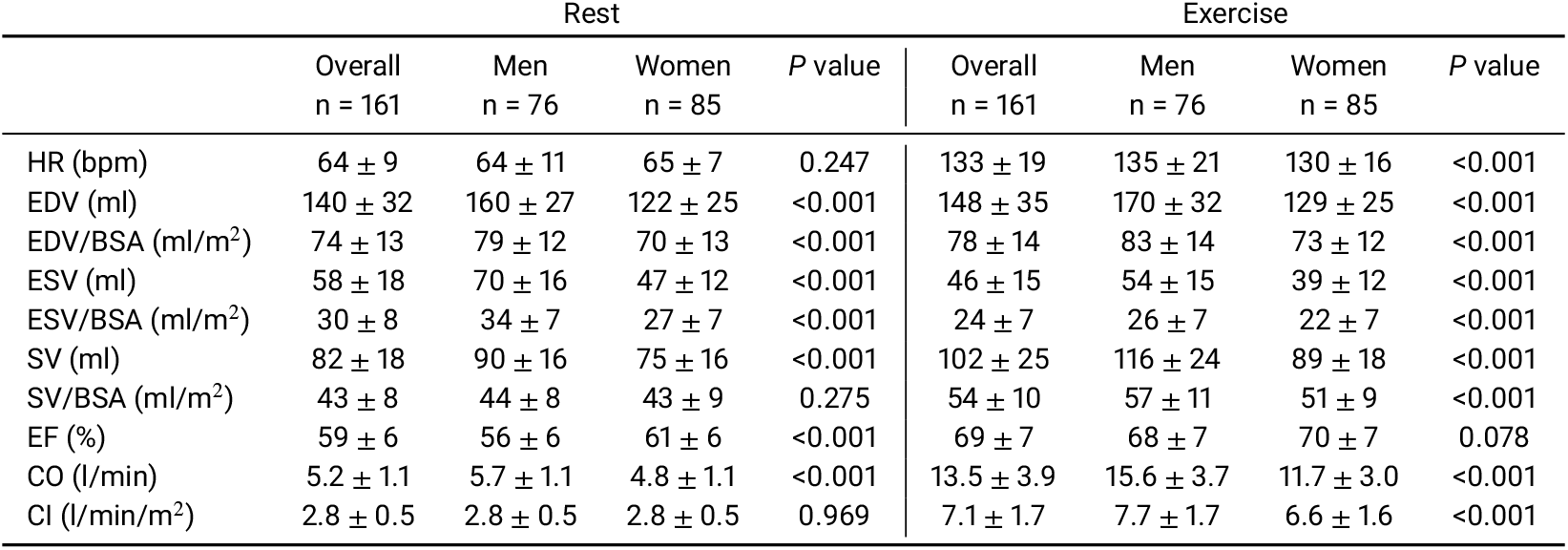
Absolute left ventricular parameters at rest and exercise in men and women. EDV, end-diastolic volume; ESV, end-systolic volume; SV, stroke volume; EF, ejection fraction; CO, cardiac output; CI, cardiac index; BSA, body surface area.

**Table S2.** Change in left ventricular parameters on exercise. EDV, end-diastolic volume; ESV, end-systolic volume; SV, stroke volume; EF, ejection fraction; CO, cardiac output; CI, cardiac index; BSA, body surface area.

|  | Overall<br>n = 161 | Men<br>n = 76 | Women<br>n = 85 | P value |
| --- | --- | --- | --- | --- |
| HR (bpm) | 68 ± 20 | 72 ± 22 | 65 ± 18 | 0.105 |
| EDV (ml) | 8 ± 20 | 10 ± 22 | 6 ± 18 | 0.12 |
| EDV/BSA (ml/m<sup>2</sup>) | 4 ± 10 | 5 ± 11 | 4 ± 10 | 0.309 |
| ESV (ml) | -12 ± 12 | -16 ± 13 | -8 ± 9 | <0.001 |
| ESV/BSA (ml/m<sup>2</sup>) | -6 ± 6 | -8 ± 7 | -4 ± 5 | <0.001 |
| SV (ml) | 20 ± 19 | 26 ± 20 | 14 ± 18 | <0.001 |
| SV/BSA (ml/m<sup>2</sup>) | 10 ± 10 | 13 ± 9 | 8 ± 10 | 0.004 |
| EF (%) | 10 ± 8 | 12 ± 7 | 8 ± 8 | 0.003 |
| CO (l/min) | 8.3 ± 3.5 | 10.0 ± 3.6 | 6.8 ± 2.6 | <0.001 |
| CI (l/min/m<sup>2</sup>) | 4.4 ± 1.7 | 4.9 ± 1.8 | 3.9 ± 1.4 | <0.001 |

**Table S3.**
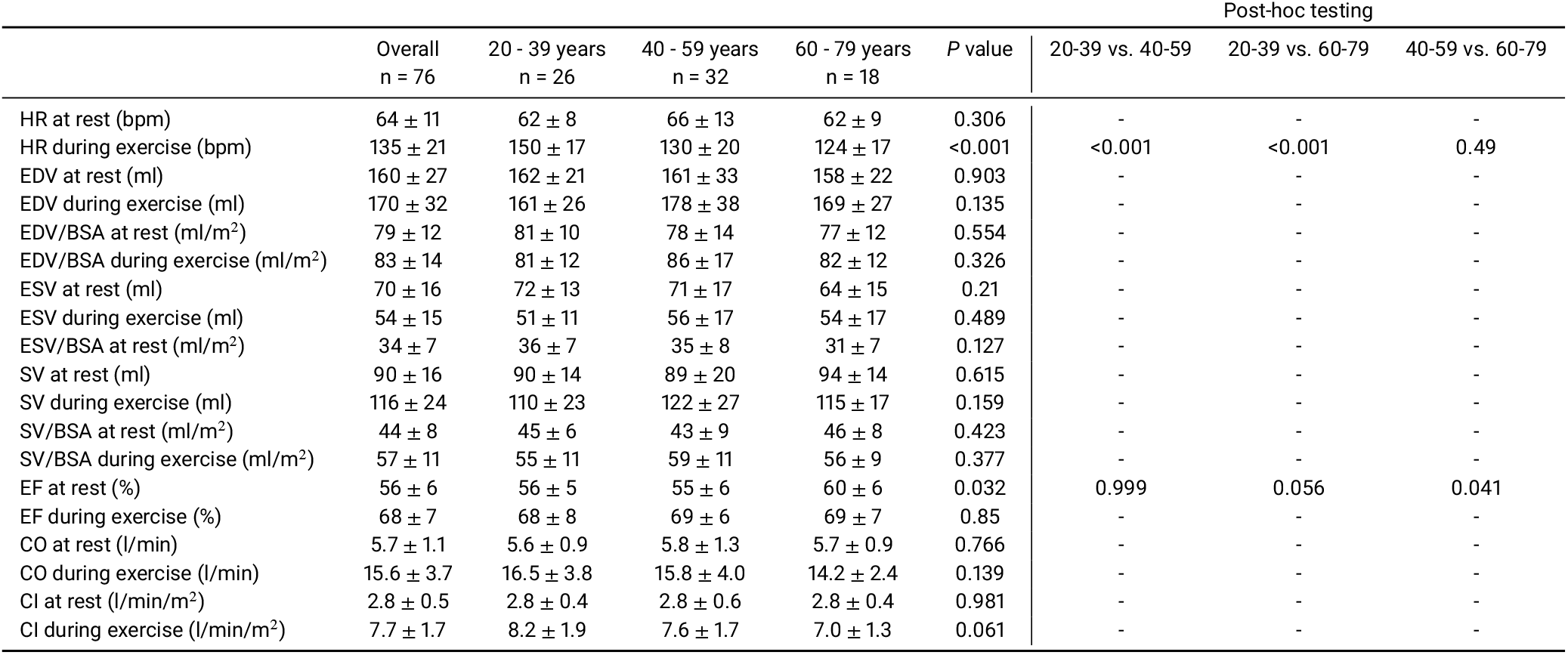
Absolute left ventricular parameters at rest and exercise in men by age group. EDV, end-diastolic volume; ESV, end-systolic volume; SV, stroke volume; EF, ejection fraction; CO, cardiac output; CI, cardiac index; BSA, body surface area; HR, heart rate.

**Table S4.** Change in left ventricular parameters on exercise in men by age group. EDV, end-diastolic volume; ESV, end-systolic volume; SV, stroke volume; EF, ejection fraction; CO, cardiac output; CI, cardiac index; BSA, body surface area; HR, heart rate.

|  | Overall<br>n = 76 | 20 - 39 years<br>n = 26 | 40 - 59 years<br>n = 32 | 60 - 79 years<br>n = 18 | P value | Post-hoc testing |  |  |
| --- | --- | --- | --- | --- | --- | --- | --- | --- |
|  |  |  |  |  |  | 20-39 vs. 40-59 | 20-39 vs. 60-79 | 40-59 vs. 60-79 |
| HR (bpm) | 72 ± 22 | 88 ± 16 | 64 ± 22 | 62 ± 17 | <0.001 | <0.001 | <0.001 | 0.932 |
| EDV (ml) | 10 ± 22 | 0 ± 24 | 18 ± 18 | 11 ± 21 | 0.007 | 0.005 | 0.194 | 0.51 |
| EDV/BSA (ml/m<sup>2</sup>) | 5 ± 11 | 0 ± 12 | 9 ± 9 | 5 ± 10 | 0.01 | 0.007 | 0.26 | 0.487 |
| ESV (ml) | -16 ± 13 | -21 ± 13 | -15 ± 12 | -11 ± 14 | 0.046 | 0.264 | 0.039 | 0.464 |
| ESV/BSA (ml/m<sup>2</sup>) | -8 ± 7 | -10 ± 6 | -7 ± 6 | -5 ± 8 | 0.042 | 0.171 | 0.042 | 0.616 |
| SV (ml) | 26 ± 20 | 20 ± 21 | 33 ± 19 | 22 ± 15 | 0.024 | 0.031 | 0.964 | 0.109 |
| SV/BSA (ml/m<sup>2</sup>) | 13 ± 9 | 10 ± 11 | 16 ± 8 | 10 ± 7 | 0.039 | 0.057 | 0.994 | 0.123 |
| EF (%) | 12 ± 7 | 12 ± 8 | 13 ± 6 | 9 ± 7 | 0.181 | - | - | - |
| CO (l/min) | 10.0 ± 3.6 | 10.9 ± 3.7 | 10.0 ± 4.0 | 8.5 ± 2.0 | 0.092 | - | - | - |
| CI (l/min/m<sup>2</sup>) | 4.9 ± 1.8 | 5.5 ± 1.9 | 4.8 ± 1.8 | 4.2 ± 1.1 | 0.059 | - | - | - |

**Table S5.** Absolute left ventricular parameters at rest and exercise in women by age group. EDV, end-diastolic volume; ESV, end-systolic volume; SV, stroke volume; EF, ejection fraction; CO, cardiac output; CI, cardiac index; BSA, body surface area; HR, heart rate.

|  | Overall<br>n = 85 | 20 - 39 years<br>n = 34 | 40 - 59 years<br>n = 35 | 60 - 79 years<br>n = 26 | P value | Post-hoc testing |  |  |
| --- | --- | --- | --- | --- | --- | --- | --- | --- |
|  |  |  |  |  |  | 20-39 vs. 40-59 | 20-39 vs. 60-79 | 40-59 vs. 60-79 |
| HR at rest (bpm) | 65 ± 7 | 66 ± 8 | 65 ± 7 | 65 ± 7 | 0.707 | - | - | - |
| HR during exercise (bpm) | 130 ± 16 | 140 ± 17 | 131 ± 13 | 121 ± 14 | <0.001 | 0.041 | <0.001 | 0.039 |
| EDV at rest (ml) | 122 ± 25 | 133 ± 26 | 122 ± 22 | 113 ± 23 | 0.013 | 0.162 | 0.009 | 0.339 |
| EDV during exercise (ml) | 129 ± 25 | 138 ± 32 | 126 ± 21 | 122 ± 18 | 0.066 | - | - | - |
| EDV/BSA at rest (ml/m<sup>2</sup>) | 70 ± 13 | 75 ± 14 | 68 ± 11 | 67 ± 13 | 0.035 | 0.067 | 0.049 | 0.952 |
| EDV/BSA during exercise (ml/m<sup>2</sup>) | 73 ± 12 | 78 ± 16 | 71 ± 11 | 73 ± 10 | 0.083 | - | - | - |
| ESV at rest (ml) | 47 ± 12 | 53 ± 14 | 46 ± 10 | 43 ± 11 | 0.01 | 0.059 | 0.009 | 0.607 |
| ESV during exercise (ml) | 39 ± 12 | 41 ± 14 | 38 ± 13 | 39 ± 10 | 0.742 | - | - | - |
| ESV/BSA at rest (ml/m<sup>2</sup>) | 27 ± 7 | 30 ± 7 | 26 ± 6 | 26 ± 6 | 0.019 | 0.031 | 0.038 | 0.996 |
| ESV/BSA during exercise (ml/m<sup>2</sup>) | 22 ± 7 | 23 ± 8 | 22 ± 7 | 23 ± 6 | 0.56 | - | - | - |
| SV at rest (ml) | 75 ± 16 | 80 ± 15 | 75 ± 17 | 70 ± 16 | 0.094 | - | - | - |
| SV during exercise (ml) | 89 ± 18 | 97 ± 24 | 88 ± 15 | 83 ± 15 | 0.027 | 0.135 | 0.023 | 0.595 |
| SV/BSA at rest (ml/m<sup>2</sup>) | 43 ± 9 | 45 ± 8 | 42 ± 8 | 41 ± 9 | 0.254 | - | - | - |
| SV/BSA during exercise (ml/m<sup>2</sup>) | 51 ± 9 | 55 ± 12 | 49 ± 7 | 49 ± 7 | 0.04 | 0.048 | 0.088 | 0.993 |
| EF at rest (%) | 61 ± 6 | 60 ± 6 | 62 ± 7 | 62 ± 6 | 0.573 | - | - | - |
| EF during exercise (%) | 70 ± 7 | 70 ± 7 | 70 ± 7 | 68 ± 7 | 0.453 | - | - | - |
| CO at rest (l/min) | 4.8 ± 1.1 | 5.2 ± 1.1 | 4.8 ± 1.0 | 4.5 ± 1.0 | 0.032 | 0.32 | 0.024 | 0.325 |
| CO during exercise (l/min) | 11.7 ± 3.0 | 13.6 ± 3.6 | 11.5 ± 2.2 | 10.1 ± 2.2 | <0.001 | 0.011 | <0.001 | 0.143 |
| CI at rest (l/min/m<sup>2</sup>) | 2.8 ± 0.5 | 3.0 ± 0.6 | 2.7 ± 0.5 | 2.6 ± 0.5 | 0.065 | - | - | - |
| CI during exercise (l/min/m<sup>2</sup>) | 6.6 ± 1.6 | 7.7 ± 2.0 | 6.4 ± 1.2 | 6.0 ± 1.2 | <0.001 | 0.005 | <0.001 | 0.49 |

**Table S6.** Change in left ventricular parameters on exercise in women by age group. EDV, end-diastolic volume; ESV, end-systolic volume; SV, stroke volume; EF, ejection fraction; CO, cardiac output; CI, cardiac index; BSA, body surface area; HR, heart rate.

|  | Overall<br>n = 85 | 20 - 39 years<br>n = 34 | 40 - 59 years<br>n = 35 | 60 - 79 years<br>n = 26 | P value | Post-hoc testing |  |  |
| --- | --- | --- | --- | --- | --- | --- | --- | --- |
|  |  |  |  |  |  | 20-39 vs. 40-59 | 20-39 vs. 60-79 | 40-59 vs. 60-79 |
| HR (bpm) | 65 ± 18 | 74 ± 18 | 66 ± 15 | 57 ± 17 | 0.002 | 0.158 | 0.001 | 0.09 |
| EDV (ml) | 6 ± 18 | 5 ± 15 | 5 ± 20 | 10 ± 17 | 0.54 | - | - | - |
| EDV/BSA (ml/m<sup>2</sup>) | 4 ± 10 | 3 ± 8 | 3 ± 11 | 6 ± 10 | 0.454 | - | - | - |
| ESV (ml) | -8 ± 9 | -12 ± 10 | -8 ± 9 | -4 ± 8 | 0.006 | 0.112 | 0.004 | 0.284 |
| ESV/BSA (ml/m<sup>2</sup>) | -4 ± 5 | -7 ± 6 | -4 ± 5 | -2 ± 5 | 0.006 | 0.098 | 0.005 | 0.334 |
| SV (ml) | 14 ± 18 | 17 ± 15 | 13 ± 19 | 14 ± 18 | 0.579 | - | - | - |
| SV/BSA (ml/m<sup>2</sup>) | 8 ± 10 | 10 ± 8 | 7 ± 10 | 8 ± 11 | 0.6 | - | - | - |
| EF (%) | 8 ± 8 | 10 ± 7 | 8 ± 8 | 6 ± 8 | 0.226 | - | - | - |
| CO (l/min) | 6.8 ± 2.6 | 8.3 ± 3.1 | 6.6 ± 2.1 | 5.7 ± 1.9 | <0.001 | 0.022 | <0.001 | 0.282 |
| CI (l/min/m<sup>2</sup>) | 3.9 ± 1.4 | 4.7 ± 1.7 | 3.7 ± 1.2 | 3.4 ± 1.1 | 0.002 | 0.018 | 0.001 | 0.519 |

**Table S7.**
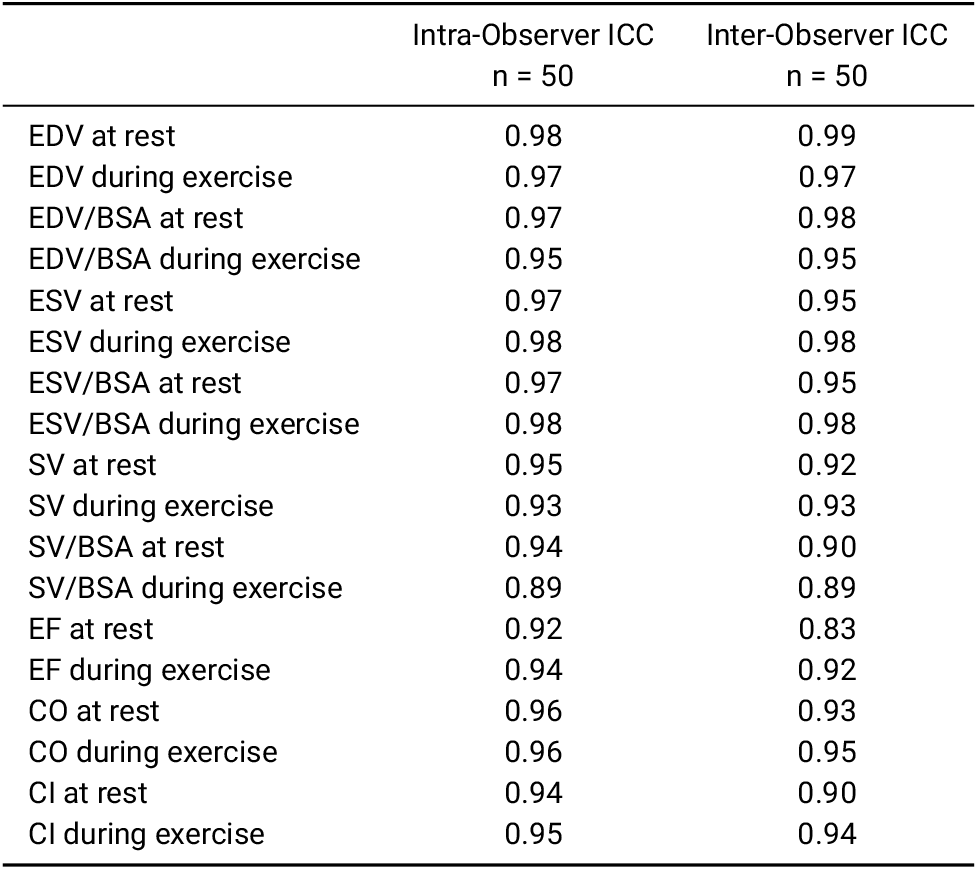
Intra- and inter-observer variability. EDV, end-diastolic volume; ESV, end-systolic volume; SV, stroke volume; EF, ejection fraction; CO, cardiac output; CI, cardiac index; BSA, body surface area.

## References

Craven TP, Tsao CW, La Gerche A, Simonetti OP, and Greenwood JP. Exercise cardiovascular magnetic resonance: development, current utility and future applications. J Cardiovasc Magn Reson. 2020;22:65.

Backhaus SJ, Lange T, George EF, Hellenkamp K, Gertz RJ, Billing M, Wachter R, Steinmetz M, Kutty S, Raaz U, Lotz J, Friede T, Uecker M, Hasenfuss G, Seidler T, and Schuster A. Exercise Stress Real-Time Cardiac Magnetic Resonance Imaging for Noninvasive Characterization of Heart Failure With Preserved Ejection Fraction: The HFpEF-Stress Trial. Circulation. 2021;143:1484–98.

Reiter C, Reiter U, Krauter C, Nizhnikava V, Greiser A, Scherr D, Schmidt A, Fuchsjager M, and Reiter G. Differences in left ventricular and left atrial function assessed during breath-holding and breathing. Eur J Radiol. 2021;141:109756.

Fletcher GF, Ades PA, Kligfield P, Arena R, Balady GJ, Bittner VA, Coke LA, Fleg JL, Forman DE, Gerber TC, Gulati M, Madan K, Rhodes J, Thompson PD, and Williams MA. Exercise standards for testing and training: a scientific statement from the American Heart Association. Circulation. 2013;128:873–934.

Kawel-Boehm N, Hetzel SJ, Ambale-Venkatesh B, Captur G, Francois CJ, Jerosch-Herold M, Salerno M, Teague SD, Valsangiacomo-Buechel E, van der Geest RJ, and Bluemke DA. Reference ranges (“normal values”) for cardiovascular magnetic resonance (CMR) in adults and children: 2020 update. J Cardiovasc Magn Reson. 2020;22:87.

Roberts AM, Ware JS, Herman DS, Schafer S, Baksi J, Bick AG, Buchan RJ, Walsh R, John S, Wilkinson S, Mazzarotto F, Felkin LE, Gong S, MacArthur JAL, Cunningham F, Flannick J, Gabriel SB, Altshuler DM, Macdonald PS, Heinig M, Keogh AM, Hayward CS, Banner NR, Pennell DJ, O’Regan DP, San TR, de Marvao A, Dawes TJW, Gulati A, Birks EJ, Yacoub MH, Radke M, Gotthardt M, Wilson JG, O’Donnell CJ, Prasad SK, Barton PJR, Fatkin D, Hubner N, Seidman JG, Seidman CE, and Cook SA. Integrated allelic, transcriptional, and phenomic dissection of the cardiac effects of titin truncations in health and disease. Sci Transl Med. 2015;7:270ra6.

Pua CJ, Bhalshankar J, Miao K, Walsh R, John S, Lim SQ, Chow K, Buchan R, Soh BY, Lio PM, Lim J, Schafer S, Lim JQ, Tan P, Whiffin N, Barton PJ, Ware JS, and Cook SA. Development of a comprehensive sequencing assay for inherited cardiac condition genes. J Cardiovasc Transl Res. 2016;9:3–11.

Schafer S, de Marvao A, Adami E, Fiedler LR, Ng B, Khin E, Rackham OJ, van Heesch S, Pua CJ, Kui M, Walsh R, Tayal U, Prasad SK, Dawes TJ, Ko NS, Sim D, Chan LL, Chin CW, Mazzarotto F, Barton PJ, Kreuchwig F, de Kleijn DP, Totman T, Biffi C, Tee N, Rueckert D, Schneider V, Faber A, Regitz-Zagrosek V, Seidman JG, Seidman CE, Linke WA, Kovalik JP, O’Regan D, Ware JS, Hubner N, and Cook SA. Titin-truncating variants affect heart function in disease cohorts and the general population. Nat Genet. 2017;49:46–53.

Lota AS, Hazebroek MR, Theotokis P, Wassall R, Salmi S, Halliday BP, Tayal U, Verdonschot J, Meena D, Owen R, et al. Genetic architecture of acute myocarditis and the overlap with inherited cardiomyopathy. Circulation. 2022;146:1123–34.

Kramer CM, Barkhausen J, Bucciarelli-Ducci C, Flamm SD, Kim RJ, and Nagel E. Standardized cardiovascular magnetic resonance imaging (CMR) protocols: 2020 update. J Cardiovasc Magn Reson. 2020;22.

Fox SM, Naughton JP, and Haskell WL. Physical activity and the prevention of coronary heart disease. Ann Clin Res. 1971;3:404–32.

Hatipoglu S, Mohiaddin RH, Gatehouse P, Alpendurada F, Baksi AJ, Izgi C, Prasad SK, Pennell DJ, and Krupickova S. Performance of artificial intelligence for biventricular cardiovascular magnetic resonance volumetric analysis in the clinical setting. Int J Cardiovasc Imaging. 2022;38:2413–24.

Bobak CA, Barr PJ, and O’Malley AJ. Estimation of an inter-rater intra-class correlation coefficient that overcomes common assumption violations in the assessment of health measurement scales. BMC Med Res Methodol. 2018;18:93.

Leiner T, Bogaert J, Friedrich MG, Mohiaddin R, Muthurangu V, Myerson S, Powell AJ, Raman SV, and Pennell DJ. SCMR Position Paper (2020) on clinical indications for cardiovascular magnetic resonance. J Cardiovasc Magn Reson. 2020;22:76.

Arai AE, Kwong RY, Salerno M, Greenwood JP, and Bucciarelli-Ducci C. Society for Cardiovascular Magnetic Resonance perspective on the 2021 AHA/ACC Chest Pain Guidelines. J Cardiovasc Magn Reson. 2022;24:8.

Raman SV, Hachamovitch R, Scandling D, Mazur W, Kwong RY, Wong TC, Schelbert EB, Moore S, Truong V, and Simonetti OP. Lower Ischemic Heart Disease Diagnostic Costs With Treadmill Stress CMR Versus SPECT: A Multicenter, Randomized Trial. JACC Cardiovasc Imaging. 2020;13:1840–2.

1. Raman SV, Dickerson JA, Mazur W, Wong TC, Schelbert EB, Min JK, Scandling D, Bartone C, Craft JT, Thavendiranathan P, Mazzaferri Jr EL, Arnold JW, Gilkeson R, and Simonetti OP. Diagnostic performance of treadmill exercise cardiac magnetic resonance: The prospective, multicenter exercise CMR’s accuracy for cardiovascular stress testing (EXACT) trial. J Am Heart Assoc. 2016;5.

Le TT, Ang BWY, Bryant JA, Chin CY, Yeo KK, Wong PEH, Ho KW, Tan JWC, Lee PT, Chin CWL, and Cook SA. Multiparametric exercise stress cardiovascular magnetic resonance in the diagnosis of coronary artery disease: the EMPIRE trial. J Cardiovasc Magn Reson. 2021;23:17.

Backhaus SJ, Lange T, George EF, Hellenkamp K, Gertz RJ, Billing M, Wachter R, Steinmetz M, Kutty S, Raaz U, Lotz J, Friede T, Uecker M, Hasenfuß G, Seidler T, and Schuster A. Exercise stress real-time cardiac magnetic resonance imaging for noninvasive characterization of heart failure with preserved ejection fraction: The HFpEF-stress trial. Circulation. 2021;143:1484–98.

Le TT, Bryant JA, Ang BWY, Pua CJ, Su B, Ho PY, Lim S, Huang W, Lee PT, Tang HC, Chin CT, Tan BY, Cook SA, and Chin CWL. The application of exercise stress cardiovascular magnetic resonance in patients with suspected dilated cardiomyopathy. J. Cardiovasc. Magn. Reson. 2020;22:10.

Claessen G, Schnell F, Bogaert J, Claeys M, Pattyn N, De Buck F, Dymarkowski S, Claus P, Carré F, Van Cleemput J, La Gerche A, and Heidbuchel H. Exercise cardiac magnetic resonance to differentiate athlete’s heart from structural heart disease. Eur. Heart J. Cardiovasc. Imaging. 2018;19:1062–70.

Jaijee S, Quinlan M, Tokarczuk P, Clemence M, Howard L, Gibbs JSR, and O’Regan DP. Exercise cardiac MRI unmasks right ventricular dysfunction in acute hypoxia and chronic pulmonary arterial hypertension. Am J Physiol Heart Circ Physiol. 2018;315:H950–H957.

Craven TP, Jex N, Chew PG, Higgins DM, Bissell MM, Brown LAE, Saunderson CED, Das A, Chowdhary A, Dall’Armellina E, Levelt E, Swoboda PP, Plein S, and Greenwood JP. Exercise cardiovascular magnetic resonance: feasibility and development of biventricular function and great vessel flow assessment, during continuous exercise accelerated by Compressed SENSE: preliminary results in healthy volunteers. Int J Cardiovasc Imaging. 2021;37:685–98.

Claessen G, Schnell F, Bogaert J, Claeys M, Pattyn N, De Buck F, Dymarkowski S, Claus P, Carre F, Van Cleemput J, La Gerche A, and Heidbuchel H. Exercise cardiac magnetic resonance to differentiate athlete’s heart from structural heart disease. Eur Heart J Cardiovasc Imaging. 2018;19:1062–70.

Wang X, Uecker M, and Feng L. Fast real-time cardiac MRI: a review of current techniques and future directions. Investig Magn Reson Imaging. 2021;25:252–65.

Beaudry RI, Samuel TJ, Wang J, Tucker WJ, Haykowsky MJ, and Nelson MD. Exercise cardiac magnetic resonance imaging: a feasibility study and meta-analysis. Am J Physiol Regul Integr Comp Physiol. 2018;315:R638–r645.

Steding-Ehrenborg K, Jablonowski R, Arvidsson PM, Carlsson M, Saltin B, and Arheden H. Moderate intensity supine exercise causes decreased cardiac volumes and increased outer volume variations: a cardiovascular magnetic resonance study. J Cardiovasc Magn Reson. 2013;15:96.

Fujimoto N, Hastings JL, Bhella PS, Shibata S, Gandhi NK, Carrick-Ranson G, Palmer D, and Levine BD. Effect of ageing on left ventricular compliance and distensibility in healthy sedentary humans. J Physiol. 2012;590:1871–80.

Bassareo PP and Crisafulli A. Gender differences in hemodynamic regulation and cardiovascular adaptations to dynamic exercise. Curr Cardiol Rev. 2020;16:65–72.

Dawes TJ, Corden B, Cotter S, de Marvao A, Walsh R, Ware JS, Cook SA, and O’Regan DP. Moderate Physical Activity in Healthy Adults Is Associated With Cardiac Remodeling. Circ Cardiovasc Imaging. 2016;9:e004712.

Morales MA, Yoon S, Fahmy A, Ghanbari F, Nakamori S, Rodriguez J, Yue J, Street JA, Herzka DA, Manning WJ, and Nezafat R. Highly accelerated free-breathing real-time myocardial tagging for exercise cardiovascular magnetic resonance. J Cardiovasc Magn Reson. 2023;25:56.

